# Integrated phenotypic and mutational approach defines EBF3-related HADD syndrome genotype-phenotype relationships

**DOI:** 10.1101/2020.12.07.20238691

**Authors:** Cole A. Deisseroth, Aarushi Nayak, Nathan D. Bliss, Vanesa Lerma, Ashley W. LeMaire, Vinodh Narayanan, Christopher Balak, Ginevra Zanni, Enza Maria Valente, Enrico Bertini, Paul J. Benke, Michael F. Wangler, Hsiao-Tuan Chao

## Abstract

Hypotonia, Ataxia, and Delayed Development syndrome is a neurodevelopmental disorder caused by heterozygous *Early B-Cell Factor 3* (*EBF3*) loss-of-function variants. Identified in 2016, the full spectrum of clinical findings and the relationship between the *EBF3* genotype and clinical outcomes has not been determined beyond its namesake features. We combined a phenotypic assessment of 33 individuals molecularly diagnosed with *EBF3* pathogenic variants with a meta-analysis of 34 previously reported individuals. The combined 62 unique individuals enabled comparative cross-sectional phenotype and genotype analysis in the largest cohort to date of affected individuals. Cardinal distinguishing features were identified that facilitate phenotypic stratification for clinical diagnosis. We developed assessment scales to ascertain individuals at risk for pathogenic *EBF3* variants, stratify the clinical severity, and connect variant-specific molecular phenotypes to clinical outcomes. Our findings show that a specific class of *EBF3* variants affecting the evolutionarily conserved Zinc Finger (ZNF) motif, which is critical for stabilizing the protein interaction with the DNA target sequence, is associated with an increased risk of persistent motor and language impairments. These findings highlight the impact of combining variant-specific molecular phenotypes with comprehensive clinical data to predict neurodevelopmental outcomes and potentially guide personalized decisions for therapeutic interventions.

## Introduction

*Early B-Cell Factor 3* (*EBF3* [MIM: 607407] HGNC: 19087) encodes a Collier/Olf/EBF (COE) transcription factor located on chromosome 10q26. COE transcription factors control gene expression via DNA methylation, nucleosome remodeling, and active chromatin modifications^1–3^, and regulate nervous system, immune system, bone, and muscle development^4–7^. Loss-of-function manipulations of *EBF3* homologs in worms (*unc-3, CeO/E*), flies (*knot, collier*), frogs (*Xcoe2*), and mice (*Ebf3*) are deleterious to survival and impair neurodevelopmental processes^8^. In mice, *Ebf3* has been shown to regulate neuronal proliferation, sub-type specification, and migration^6,9–11^. Furthermore, cellular studies revealed that *EBF3* binds to the promoters of many genes associated with neurodevelopmental and autism spectrum disorders^12^, suggesting that EBF3 dysfunction could result in a wider phenotypic spectrum than currently recognized.

Over 30 years ago, genomic deletions containing *EBF3* were identified in the 10q26-deletion syndrome, which is characterized by delayed development, hypotonia, ataxia, growth problems, neuropsychiatric comorbidities, and genitourinary abnormalities [MIM: 609625]^13–17^. However, the principal genetic alterations mediating pathogenesis in 10q26-deletion syndrome was unknown until recently. In 2016, *de novo EBF3* variants and a heterozygous *Ebf3* single gene deletion were discovered to cause the Hypotonia, Ataxia, and Delayed Development Syndrome (HADDS [MIM: 617330]), which is characterized by the namesake features and includes impaired expressive language, neuropsychiatric comorbidities including autism, and genitourinary abnormalities^12,18–20^. The strong overlap in features between HADDS and 10q26-deletion syndrome suggests that *EBF3* haploinsufficiency may be the principal pathogenic factor in 10q26-deletion syndrome.

Although the discovery of a monogenic disorder associated with pathogenic *EBF3* variants is fairly recent, there is increasing evidence that *EBF3*-related disorders may be more common than previously recognized because of a wide range of clinical severity^20–25^. This is consistent with a prior statistical model of *de novo* variants for autism spectrum disorder (ASD) and developmental and intellectual disability (DD/ID) that identified *EBF3* as one of ∼1,000 genes with significant enrichment of *de novo* loss-of-function variants in affected individuals^26^. Therefore, expanding our understanding of the HADDS phenotypic spectrum and co-morbid conditions will advance diagnosis, facilitate prognosis, and improve neurodevelopmental outcomes for individuals with HADDS, 10q26-deletion syndrome, and other prevalent neurodevelopmental disorders with similar features.

A common challenge faced with newly recognized or relatively rare neurodevelopmental disorders is that incomplete knowledge about distinguishing features limits the stratification of affected individuals for traditional phenotypically driven diagnostics. To expand our understanding of the HADDS phenotypic spectrum and identify individuals at risk for pathogenic *EBF3* variants, we conducted a comprehensive phenotypic and mutational analysis of 33 individuals diagnosed with HADDS and correlated the findings with a literature meta-analysis for a combined assessment of 62 unique individuals. Our findings expand our understanding of the HADDS phenotypic spectrum and reveal that *EBF3* variant type and location are associated with quantifiable clinical severity.

## MATERIALS AND METHODS

BCM investigators systematically evaluated 22 individuals in person at Texas Children’s Hospital and 11 through medical record reviews for a total of 33 individuals. Individuals were enrolled in a study approved by the Institutional Review Board of Baylor College of Medicine (BCM), following its ethical standards. Written and informed consent was obtained from all individuals before the study was conducted. All procedures followed national standards. All data were de-identified. Three individuals opted out of publishing genomic information. We compared our cohort to a meta-analysis of nine studies containing a total of 34 individuals with pathogenic *EBF3* variants^12,18–25^. Five of the individuals enrolled in our study were previously reported. Probands 7, 15, 19, 25, and 26 from our study (**Table S1**) are the same as meta-analysis probands M-11, M-1, M-25, M-31, and M-32, respectively (**Table S2**). We assessed a total of 62 unique individuals and compared demographics, clinical features, and molecular findings in our cohort to the meta-analysis (**Tables S1-S5**). Variant nomenclature is according to gene transcript (GenBank: NM_001005463.2, GRCh37). Odds ratios (OR) = AD/BC, where A is the number of probands from our cohort with the feature, B is the number of probands from our cohort without the feature, C is the number of probands from the meta-analysis with the feature, and D is the number of probands from the meta-analysis without the feature. 95% confidence intervals (CI) were calculated for a sample size *n* and proportion *p*:

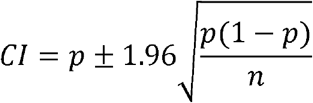

For height, weight, and occipito-frontal circumference (OFC) measurements, we calculated LMS (lambda-mu-sigma)-based Z-scores^27^, where for a given L (degree of skewness), M (median), and S (variation coefficient) values, and an experimental value X, the z-score is = [(X/M)^L^ – 1]/(LS). L, M, and S values for height and weight based on sex and age (birth to 20 years) are from the Centers for Disease Control and Prevention (CDC)^28^. For OFC, the L, M, and S values were taken from the World Health Organization (WHO, birth to five years)^29^. Delayed development was determined by failure to achieve developmental milestones within the CDC’s standard age range (**Tables S4, S5**)^30^.

### HADDS Developmental and Behavioral Scale (HADDS-DBS)

The HADDS-DBS assesses for seven major and 14 minor features (**Table 1**). Major features include decreased pain sensitivity (HP: 0007328), weak/absent cry (HP: 0001612), motor delay (HP: 0001270), hypotonia (HP: 0001252), speech delay (HP: 0000750), ataxia (HP: 0001251), and hypomimia (HP: 0000338). Minor features include poor eye contact, strabismus, repetitive scratching in any location, repetitive scratching on back or neck, attraction to lights, aversion to crowds, aversion to loud noises, constipation, frontal bossing, recurrent urinary tract infections, feeding difficulties, cerebellar anomalies, gait ataxia, and short stature. Based on the number of diagnostic features identified, the individuals are stratified into four tiers (**Table S6)**. Higher-numbered tiers indicate a higher risk for a pathogenic *EBF3* variant.

Positive (PPV) and negative (NPV) predictive values were calculated by combining the HADDS-DBS tier assignments with the molecularly confirmed genetic diagnoses. True positives (TP) are individuals with HADDS and a Tier-3 or -4 assignment. False positives (FP) are individuals with non-*EBF3* related genetic syndromes and a Tier-3 or -4 assignment. True negatives (TN) are individuals with non-*EBF3* related genetic syndromes and a Tier-1 or -2 assignment. False negatives (FN) are individuals with HADDS and a Tier-1 or -2 assignment. Sensitivity = TP/(TP+FN). Specificity = TN/(TN+FP). PPV = TP/(TP+FP). NPV = TN/(TN+FN).

For each diagnostic tier, OR = AD/BC, where A is the number of probands from our cohort at the specified tier, B is the number of probands from our cohort not at the specified tier, C is the number of probands from the meta-analysis at the specified tier, and D is the number of probands from the meta-analysis not at the specified tier. Statistically significant differences at the 0.05 level were determined by one-way ANOVA with Tukey’s post-hoc analysis.

### HADDS Developmental Delay Severity Scale (HADDS-DDSS)

The four-tier HADDS-DDSS assesses for delayed development and neuroanatomical findings in individuals at or greater than three years of age (**Table S7**). Higher-numbered tiers indicates increasingly delayed development, persistent limitations in motor and language abilities, and neuroanatomical alterations. Tier 1 includes individuals who achieved first word (12 months) and independent walking (18 months) milestones appropriately for age and without regression. Tier 2 includes individuals who achieved independent walking and first word milestones by three years of age. Tier 3 includes individuals who achieved independent walking or first word by three years of age with subsequent loss of the acquired skills or did not achieve the milestones by three years of age. Tier 4 includes individuals with neither speech nor independent walking achieved after three years of age. If information for the developmental milestones is limited, then the presence or absence of cerebellar anomalies is considered a secondary factor. If either motor or language developmental milestone is unavailable and brain imaging results are not available, then the proband is excluded from the analysis. Statistically significant differences at the 0.05 level were determined by one-way ANOVA with Tukey’s post-hoc analysis. OR for each severity tier were calculated using the same formula as the HADDS-DBS.

## RESULTS

### General features, demographics, and inheritance in HADDS

Demographics, growth parameters, genotype, and phenotype findings in our cohort were evaluated with a comparative literature meta-analysis (**Fig.1A-D, Table S3**). Growth parameters reveal below-average height and weight for individuals with HADDS (**Table S3**). However, head circumference both at birth and recent assessment remained within average parameters.

**Figure 1:**
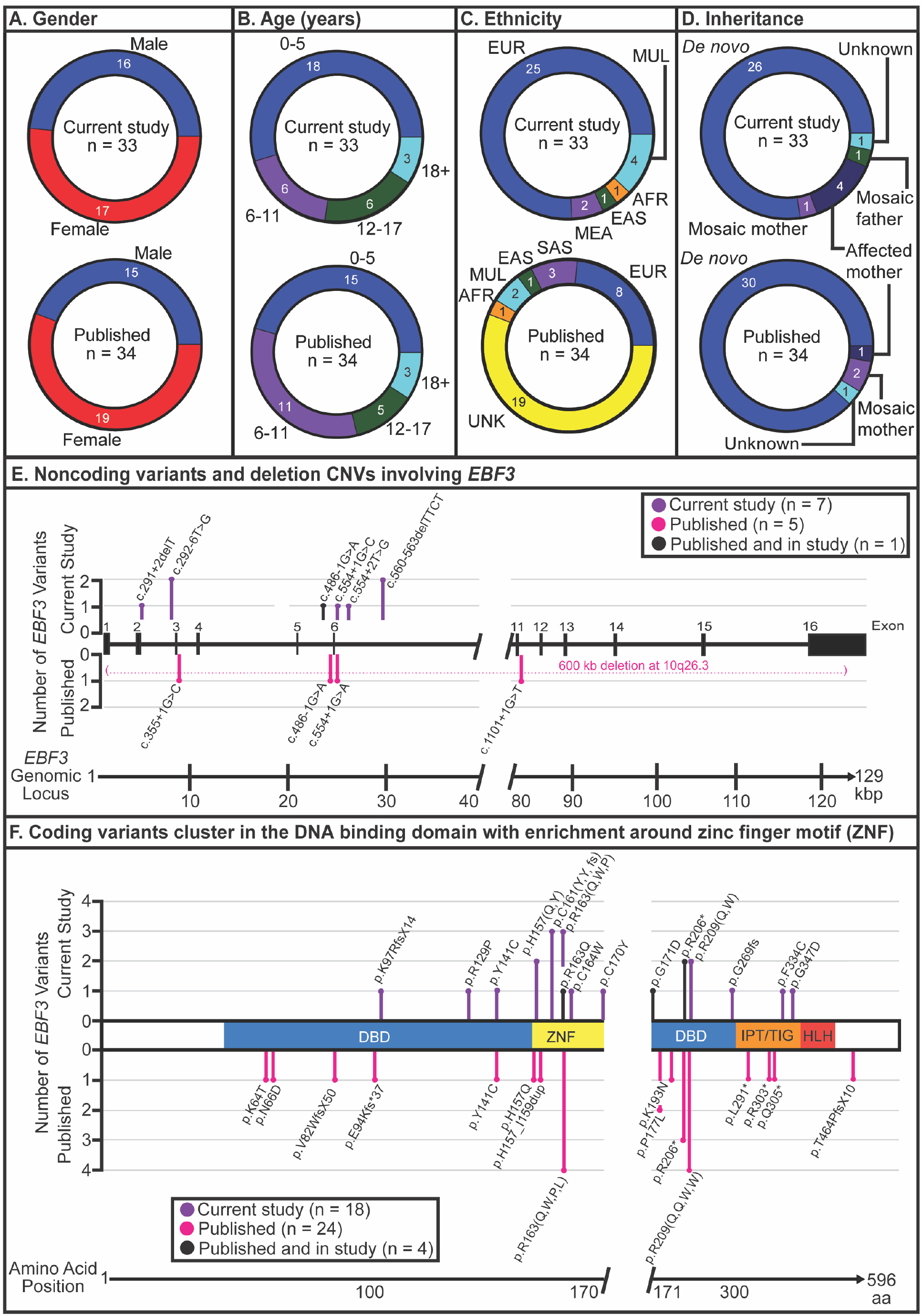
Demographics and molecular findings in *EBF3*-related HADDS. Comparison of demographic and *EBF3* variant findings from 33 individuals in this study and 34 individuals in the meta-analysis. A total of 62 unique individuals were evaluated. **(A)** Gender distribution was equivalent between our cohort and the meta-analysis. **(B)** The molecularly confirmed diagnosis of *EBF3*-related HADDS occurred predominantly in the pediatric population, with more than 90% of affected individuals identified at less than 18 years old. **(C)** Ethnicity distribution in our study compared to meta-analysis. UNK = Unknown, EUR = European, AFR = African, EAS = East Asian, SAS = South Asian, MEA = Middle Eastern, MUL = Multiethnic. **(D)** Inheritance pattern for the majority of identified *EBF3* pathogenic variants was *de novo*. **(E, F)** There are 13 individuals with noncoding *EBF3* variants and genomic deletions (**E**) and 46 individuals with coding variants (**F**). Published variants are shown in magenta, variants unique to this study are in purple, and variants shared between our cohort and the literature are in black. DBD is the DNA binding domain, ZNF is the zinc finger motif, IPT/TIG stands for immunoglobin-like, plexins, transcription factors/transcription factor immunoglobin, fold structure, and HLH is the helix-loop-helix region.

Gender distribution was equivalent between our cohort and the meta-analysis (OR=1.19) (**Fig.1A**). More than 90% of affected individuals were identified at <18 years old (**Fig.1B**). Self-reported ethnicity in our study included European (76%), African (3%), East Asian (3%), Middle Eastern (6%), and multiethnic (12%) descent. Ethnicity was reported in <45% of individuals in the meta-analysis, but there was overall a similar distribution (**Fig.1C**).

The inheritance pattern for the majority of *EBF3* pathogenic variants was *de novo* in 79% of our study and 88% of the meta-analysis (OR=0.50, **Fig.1D**). Mendelian inheritance of a pathogenic *EBF3* variant was previously reported in one affected mother and son pair^23^, and inheritance of a pathogenic *EBF3* variant from an unaffected parent with mosaicism was reported in an unaffected mother and two affected children^12^. In our study, we identified Mendelian inheritance in four affected individuals from three unrelated families (OR=4.55) and mosaicism in the parents of two affected and unrelated individuals (OR=1.03) (**Fig.1D**).

### Pathogenic *EBF3* variants and genomic deletions

We identified 13 individuals with noncoding *EBF3* variants and genomic deletions (**Fig.1E**) and 46 individuals with coding variants (**Fig.1F**). The majority of *EBF3* variants cluster within the N-terminal DNA binding domain with 27% of pathogenic variants located within five amino acids of the zinc finger motif (ZNF, amino acid 157-170, OR=2.2), a region critical for stabilizing the interaction of EBF3 with the DNA target^31^. A prior *in vitro* study of the paralogous EBF1 showed that disrupting conserved residues in the ZNF, including p.Arg163, abolished DNA-binding activity^31^. Intriguingly, we identified the most frequent recurrent *EBF3* variants affected the p.Arg163 codon in eight unrelated individuals with p.Arg163Gln (c.488G>A) in three, p.Arg163Pro (c.488G>C) in two, p.Arg163Trp (c.487C>T) in two, and p.Arg163Leu (c.488G>T) in one individual. Other recurrent variants affected the p.Arg209 codon in six, p.Arg206 codon in five, p.Cys161 codon in three, and p.His157 codon in three individuals. The recurrent variants are likely due to the position of the affected nucleotides in CpG-dinucleotide islands, which are mutational hotspots underlying over one-third of *de novo* missense variants associated with human diseases^32,33^. Other potential mechanisms include the selfish spermatogonial selection process^34^, the error-prone replication hypothesis^35^, or a selection bias for similar phenotypes.

### Common neurologic features in HADDS

Prevalent neurologic features identified in the cohort include delayed development, motor incoordination, perturbed expressive language, altered sensory processing, neuropsychiatric comorbidities, and cerebellar alterations. Consistent between our cohort and the meta-analysis, we found that more than 95% of affected individuals had delayed motor development and more than 80% had delayed expressive language development (OR=3.32, **Fig.2A**). Intriguingly, we found discordance between expressive and receptive language development with 36% (CI[16, 56]) of individuals diagnosed with HADDS able to follow one-step verbal commands on time and 37% (CI[15, 58]) able to follow two-step verbal commands on time. This finding suggests that individuals with HADDS have a greater prevalence of delayed expressive language development, potentially related to motor incoordination and speech apraxia due to cerebellar dysfunction^36,37^, but maintain age appropriate receptive language development. Another common language finding was dysarthria (30%, CI[15, 46]), which was identified in a similar frequency in the meta-analysis (OR=1.68, **Fig.2B**). We also identified speech apraxia in 27% (CI[12, 42], OR=6.00, **Fig.2B**) and weak or absent cry in 97% (CI[91, 103]) of affected individuals in our study, a higher prevalence than previously reported (OR 512, **Fig.2B**).

**Figure 2:**
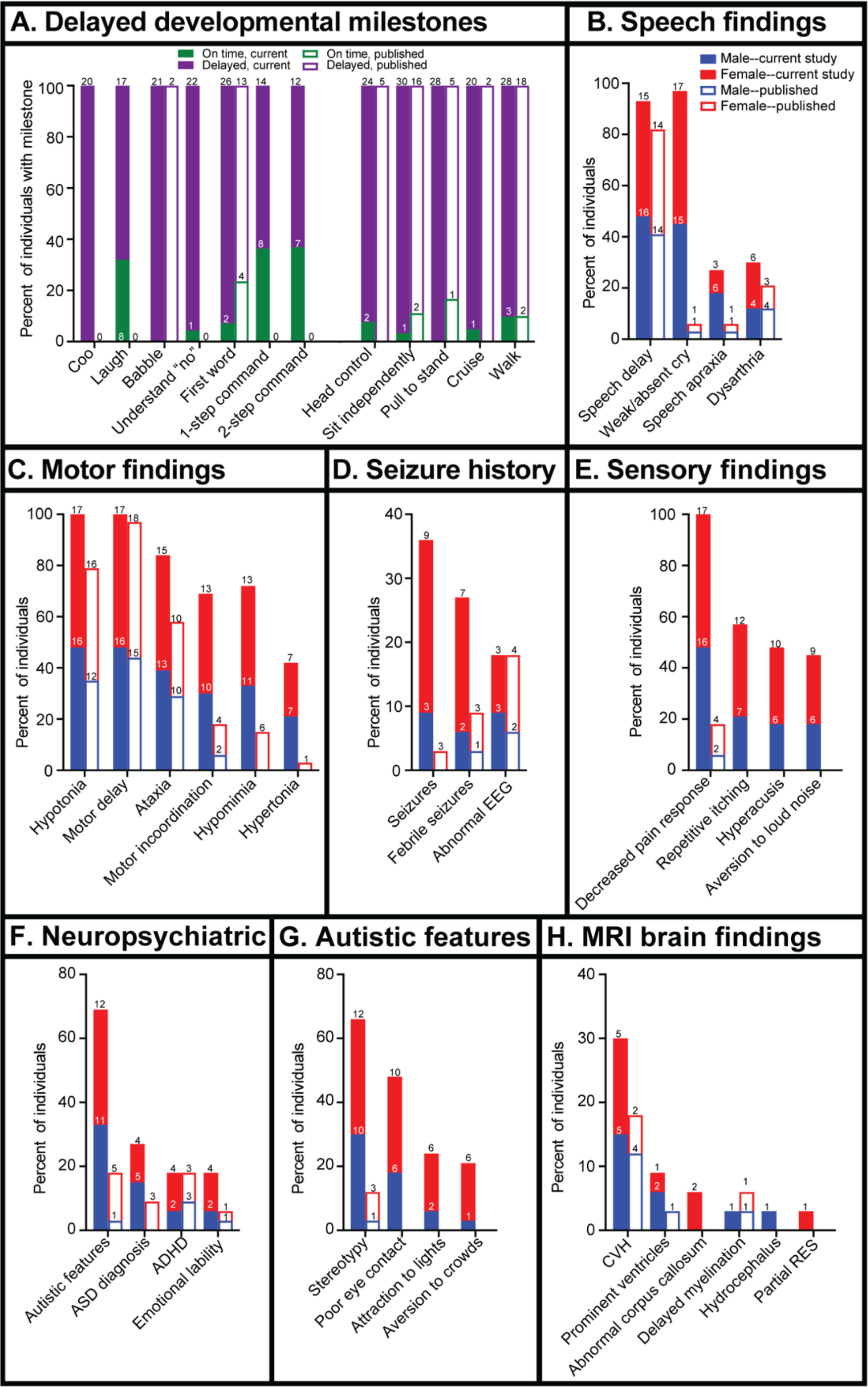
Frequency of neurological features in *EBF3*-related HADDS. From the combined analysis of 62 unique individuals, we identified distinct common neurologic features associated with pathogenic *EBF3* variants. Findings from our cohort are shown in solid bars and findings from the meta-analysis are shown in the outlined bars. Data shown as percent of individuals with the feature. Sample sizes are shown above each bar. Females are shown in red and males in blue. **(A)** Percent of individuals reporting language and gross motor developmental milestones as either on time (green) or delayed (purple) reveal prevalent motor and language delays. **(B)** Other language findings in our cohort include speech delay, weak or absent crying during infancy, speech apraxia, and dysarthria. **(C)** Frequent motor findings observed in affected individuals include hypotonia, motor delay, ataxia, motor incoordination, hypomimia, and hypertonia. **(D)** A risk of seizures, febrile seizures, and abnormal EEG findings are observed in our cohort. **(E)** Sensory alterations are prevalent in HADDS with frequent findings of decreased pain response, repetitive itching behaviors, hyperacusis, and aversion to loud noise. **(F)** Prevalent neuropsychiatric comorbidities in HADDS include autistic features, autism, attention deficit and hyperactivity disorder (ADHD), and emotional lability. **(G)** Common autistic features identified in our study include stereotypies, poor eye contact, attraction to bright lights, and aversion to crowded environments. **(H)** Neuroanatomical findings observed in our HADDS cohort include cerebellar-vermian hypoplasia (CVH), prominent ventricles, abnormal corpus callosum, delayed myelination, hydrocephalus, and partial rhomboencephalosynapsis (RES).

Consistent with the namesake features, we found a high prevalence of hypotonia (100%) and ataxia (84%, CI[73, 97], OR=3.92) in our cohort (**Fig.2C**). The phenotypic analysis in our cohort also revealed a high frequency of motor incoordination (70%, CI[54, 85], OR=10.73), hypomimia (73%, CI[57, 88], OR=12.44), and hypertonia (42%, CI[26, 59], OR=24.32), but these features were reported in less than 20% of affected individuals in the meta-analysis (**Fig.2C**). Additional neurologic findings include an increased risk of seizures (36%, CI[20, 53], OR=18.86), febrile seizures (27%, CI[12, 42], OR=3.875), and abnormalities on electroencephalogram (EEG) recordings (18%, CI[5, 31] OR=1.04) (**Fig.2D**). Intriguingly, we found that altered sensory processing is a cardinal feature of HADDS. All individuals in our study had chronic decreased pain response, but a smaller prevalence was identified in the meta-analysis (18%, CI[5, 30]), potentially due to incomplete ascertainment (**Fig.2E**). Other sensory findings in our cohort include repetitive scratching behaviors (58%, CI[41, 74]), hyperacusis (48%, CI[31, 66]), and discomfort with loud noises (45%, CI [28, 62]) (**Fig.2E**), none of which were previously reported.

Neuropsychiatric comorbidities are commonly observed in affected individuals with HADDS. We found that these features occur at a higher frequency than previously reported. Prevalent neuropsychiatric comorbidities in HADDS include autistic features (70%, CI [54, 85], OR=10.73), formal autism diagnosis (27%, CI [12, 42], OR=8.82), attention deficit and hyperactivity disorder (ADHD, 18%, CI [5, 31], OR=1.03), and emotional lability (18%, CI [5, 31], OR=3.56) (**Fig.2F**). Common autistic features identified in our study include stereotypies (67%, CI [51, 83], OR=15.0), poor eye contact (48%, CI [31, 66]), attraction to lights (24%, CI [10, 39]), and aversion to crowds (21%, CI [7, 35]) (**Fig.2G**). In contrast, the meta-analysis identified autistic features, autism, or autism spectrum disorder in 8-18% of individuals (**Fig.2F**).

Consistent with the neurodevelopmental impairments, neuroimaging studies in our cohort revealed altered cerebellar structure in 33% (CI [17, 49]) of individuals, compared to 24% (CI [9, 38]) in the meta-analysis (OR=1.63, **Fig.2H**). Cerebellar vermian hypoplasia was observed in 30% (CI [15, 46]) of individuals in our study, compared to 18% (CI [5, 30]) in the meta-analysis (OR=2.03), and is the most common neuroanatomical finding. Less frequent findings in our cohort include prominent ventricles (9%, CI [-1, 19]), abnormal corpus callosum (two individuals), delayed myelination (one individual), hydrocephalus (one individual), and partial rhomboencephalosynapsis (one individual) (**Fig.2H**).

### Non-neurological findings in HADDS

Other features commonly seen in HADDS include craniofacial, gastrointestinal, ophthalmologic, genitourinary, and musculoskeletal involvement. In our study, 88% (CI[77, 99]) of individuals had at least one craniofacial feature (OR=0.97). Recurrent features included deep-set eyes or prominent forehead (64%, CI[47, 80]), ear abnormalities (39%, CI[22, 56]), mouth anomalies (42%, CI[26, 59]), straight eyebrows (27%, CI[12, 42]), tall forehead (21%, CI[7, 35]), long face (18%, CI[5, 31]), pointed chin (21%, CI[7, 35]), myopathic facies (21%, CI[7, 35]), triangular facies (9%, CI[-1, 19]), anteverted nares (9%, C [-1, 19]), and short, broad chin (6%, CI[-2, 14]) (**Fig.3A**). Ophthalmologic findings also occur frequently in HADDS with strabismus reported in 88% (CI[77, 99]) of individuals in our cohort compared to 62% (CI[45, 78]) in the meta-analysis (OR=4.49) and nystagmus reported in 6% (CI[2, 14], OR=1.03) of individuals in both our cohort and the meta-analysis (**Fig.3B**).

**Figure 3:**
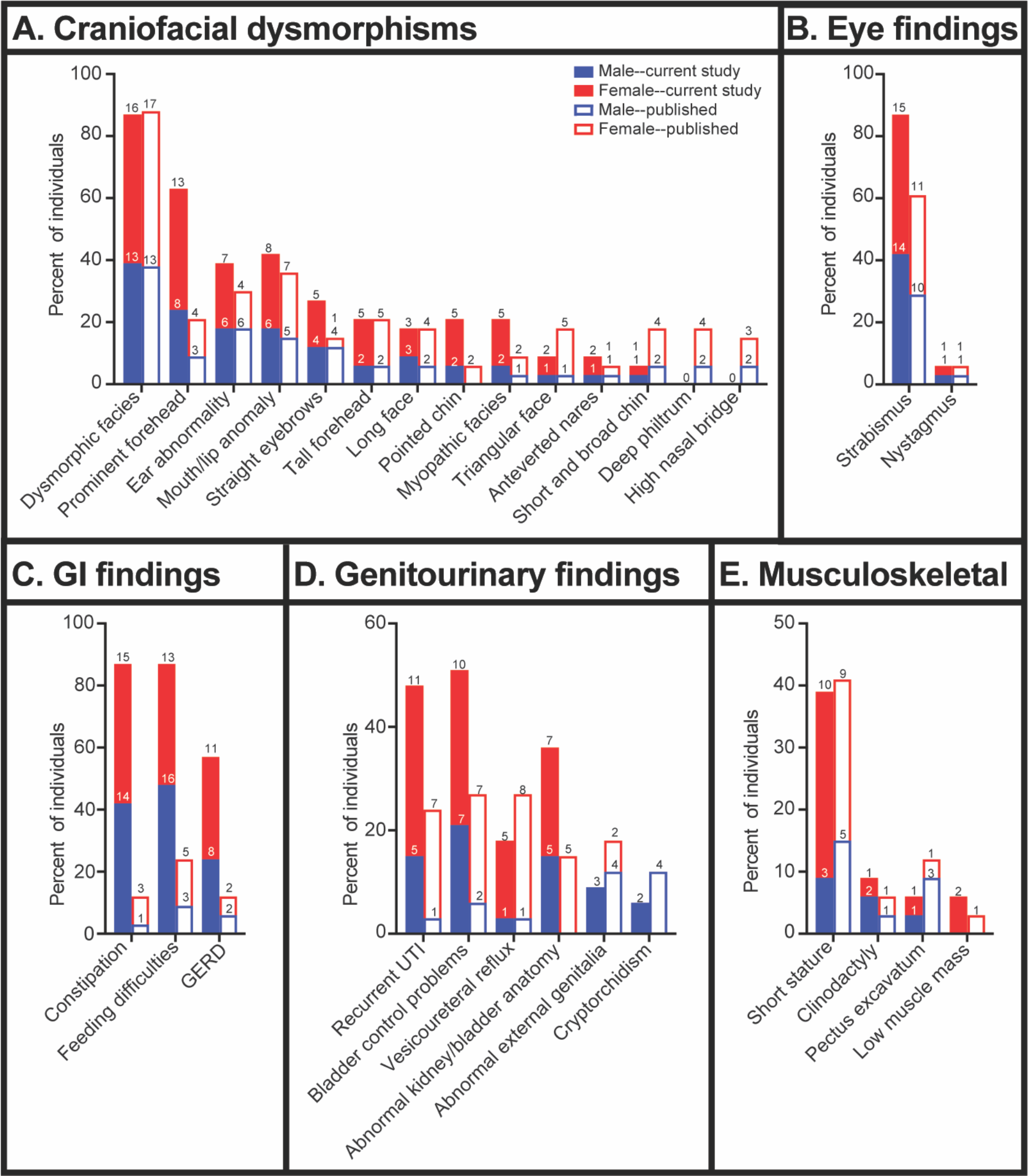
Frequency of non-neurological features in *EBF3*-related HADDS. Other features commonly seen in individuals with HADDS include craniofacial, gastrointestinal, ophthalmologic, genitourinary, and musculoskeletal involvement. Findings from the current study are shown in solid bars and findings from the meta-analysis are shown in the outlined bars. Data is shown as a percentage of individuals with the feature. Sample sizes are shown above each bar. Females are shown in red and males in blue. **(A)** Craniofacial dysmorphisms are frequently observed with 84% of affected individuals in our cohort found to have at least one craniofacial finding. **(B)** Strabismus is the most common ophthalmology finding in individuals with HADDS. Nystagmus was reported in a subset of individuals. **(C)** Constipation, feeding difficulties, and gastroesophageal reflux disease (GERD) are common gastrointestinal (GI) findings identified in affected individuals. **(D)** Genitourinary complications are frequently observed in both genders with recurrent urinary tract infections (UTIs) and bladder control problems as the most prevalent findings. Other genitourinary findings include vesicoureteral reflux (VUR), abnormal bladder and/or kidney anatomy, abnormal external genitalia, and cryptorchidism **(E)** Musculoskeletal findings include short stature, clinodactyly, pectus excavatum, and low muscle mass.

In our cohort, we identified a high frequency of constipation (88%, CI[77, 99], OR=54.38), feeding difficulties (88%, CI[77, 99], OR=24) and gastroesophageal reflux (58%, CI[41, 74], OR=10), compared to meta-analysis frequencies of 12-21% for these features (**Fig.3C**). Genitourinary complications are frequently observed in both males and females. In our cohort, we found that recurrent urinary tract infections (UTIs) occurred in 48% (CI[31, 66]) of individuals compared to 24% (CI[9, 38]) in the meta-analysis (OR=3.06), with a higher prevalence in females (**Fig.3D**). Anatomical findings in our study included abnormal kidney or bladder anatomy (36%, CI[20, 53], OR=3.31), vesicoureteric reflux (18%, CI[5, 31], OR=0.62), abnormal external genitalia (9%, CI[-1, 19], OR=0.47), and cryptorchidism (6%, CI[-2, 14], OR=0.48) (**Fig.3D**), which had a similar frequency in the meta-analysis. Female reproductive organ anomalies were seen in 6% of the meta-analysis (**Table S2**) but were not observed in our cohort (**Table S1**). The most prevalent musculoskeletal finding was short stature, which we identified in 39% (CI[23, 56]) of individuals in our cohort and was similarly reported in 41% (CI[24, 58] of the meta-analysis (OR=0.71). Other musculoskeletal dysmorphisms occurred in less than 10% of individuals in our cohort and in the meta-analysis (OR≤3). Dysmorphisms included clinodactyly (9%, CI[-1,19]), pectus excavatum (6%, CI[-2,14]), and low muscle mass (6%, CI[-2,14]) (**Fig.3E**). Short digits were also reported in two individuals in the literature (**Table S2**).

### Diagnostic utility of the HADDS developmental and behavioral scale

To aid clinical recognition and expedited diagnosis of HADDS, we developed a diagnostic assessment scale (HADDS-DBS) to identify individuals “at risk” for pathogenic *EBF3* variants and prioritize *EBF3*-specific gene testing. We designed the HADDS-DBS to address the challenges of assessing children in various age groups and developed the scale for use by caregivers and healthcare providers. The HADDS-DBS can be used to screen for *EBF3* pathogenic variant “risk” in children as young as 15-months old and takes the form of a symptom questionnaire comprised of 21 clinical features (**Table 1**). Based on the number of major and minor clinical features identified in the questionnaire, the HADDS-DBS assigns one of four tiers based on increasing risk of carrying a pathogenic *EBF3* variant with recommendations for molecular testing (**Table S6**). When applied to our cohort, the HADDS-DBS identified 30 individuals as Tier-4, two individuals as Tier-3, and one individual as Tier-2 (**Fig.4A, Table S8**). When applied to the 34 individuals in the meta-analysis, the HADDS-DBS identified three individuals as Tier-4 (OR=103), seven individuals as Tier-3 (OR=0.25), 12 individuals as Tier-2 (OR=0.06), and 12 individuals as Tier-1 (**Fig.4A, Table S9**). These differences in phenotypic stratification between our cohort and the meta-analysis reflects the variability in phenotypic ascertainment across different clinical sites and highlights the value of comprehensive clinical phenotyping (**Fig.4A**, *p*<0.001, F=102.04).

**Figure 4:**
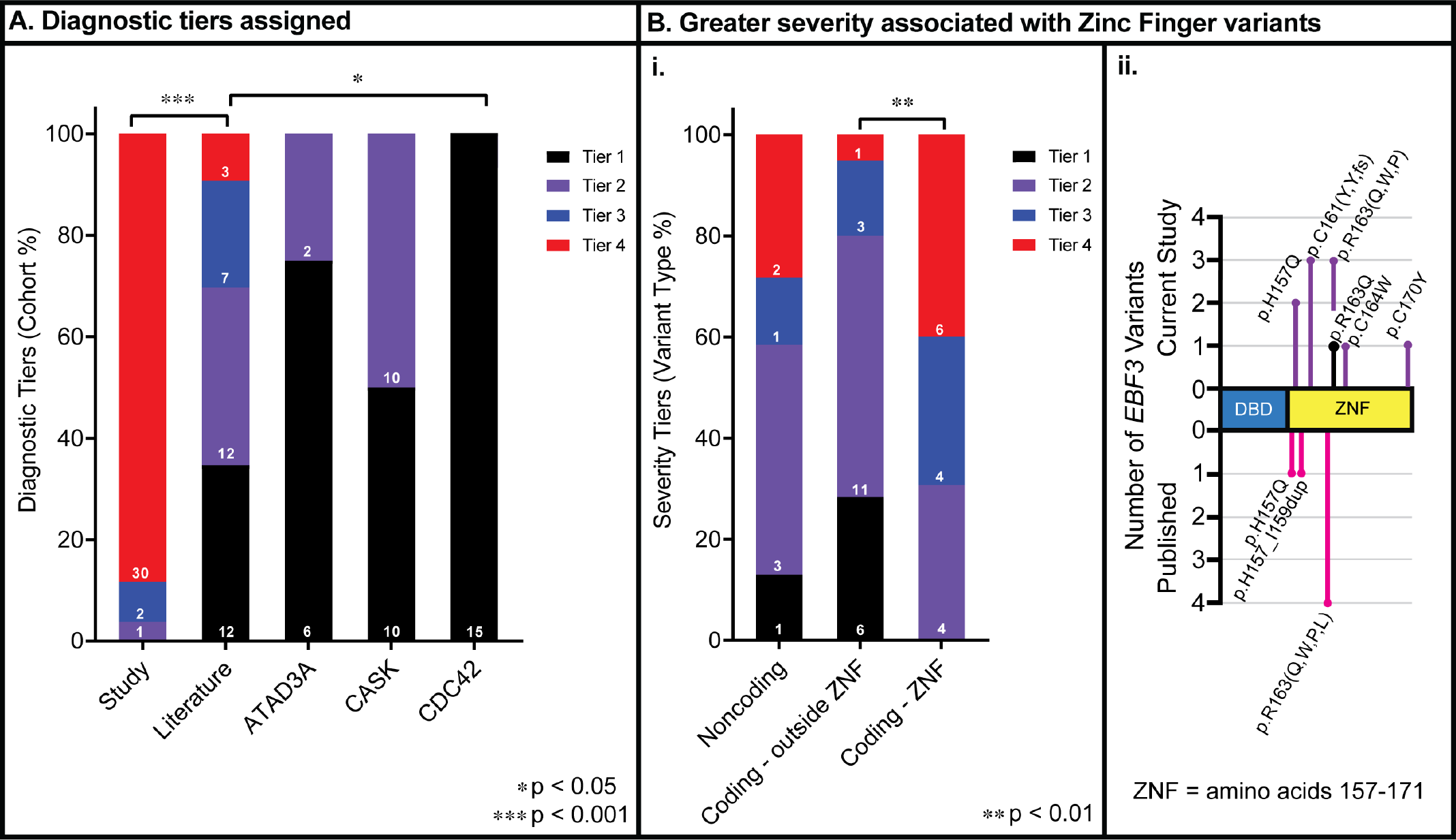
Diagnostic assessment scale and genotype-phenotype mapping in *EBF3*-related HADDS. The HADDS-DBS is a symptom questionnaire designed to identify individuals at risk for pathogenic *EBF3* variants. **(A)** The HADDS-DBS diagnostic scale was applied to individuals with molecularly confirmed HADDS enrolled in our study and previously reported in the literature. The majority of affected individuals enrolled in our study are classified as Tier-4, whereas the majority of affected individuals in the meta-analysis are classified as Tiers-2 and -3. For the five probands in our study who were previously published, we used the classification etermined from phenotypic information collected for the study. When applied to three non-*EBF3* related disorders [MIM: 617183, 300749, 616737], the HADDS-DBS primarily classified the reported individuals as Tier-1. Data is shown as a percentage of each cohort classified in Tiers-1 through -4. Sample sizes are shown above each bar. Tier-1 is shown in black, Tier-2 is shown in purple, Tier-3 is shown in blue, and Tier-4 is shown in red. Statistically significant differences were determined by one-way ANOVA with Tukey’s post-hoc analysis. Significance shown as * *p*<0.05 and *** *p*<0.001. **(B)** The HADDS-DDSS is a symptom assessment scale designed to determine the severity of neurological limitations. The scale is based on readily ascertained developmental milestones (age of first word and age of independent ambulation) and the presence or absence of MRI brain findings. **(i)** The HADDS-DDSS tier assignment is examined relative to the *EBF3* variant type and location relative to the ZNF motif for genotype-phenotype mapping. A significant correlation was identified between symptom severity and coding variants within the ZNF motif (amino acids 157-170) and the flanking five amino acids (total range 152-175). Data is shown as the percentage of each cohort classified in Tiers-1 through -4. Sample sizes are shown above each bar. Tier-1 is in black, Tier-2 is in purple, Tier-3 is in blue, and Tier-4 is in red. Statistically significant differences were determined by one-way ANOVA with Tukey’s post-hoc analysis. Significance shown as ** p<0.01. **(ii)** Illustration shows the distribution of *EBF3* missense variants within five amino acids flanking the ZNF motif.

**Table 1:**
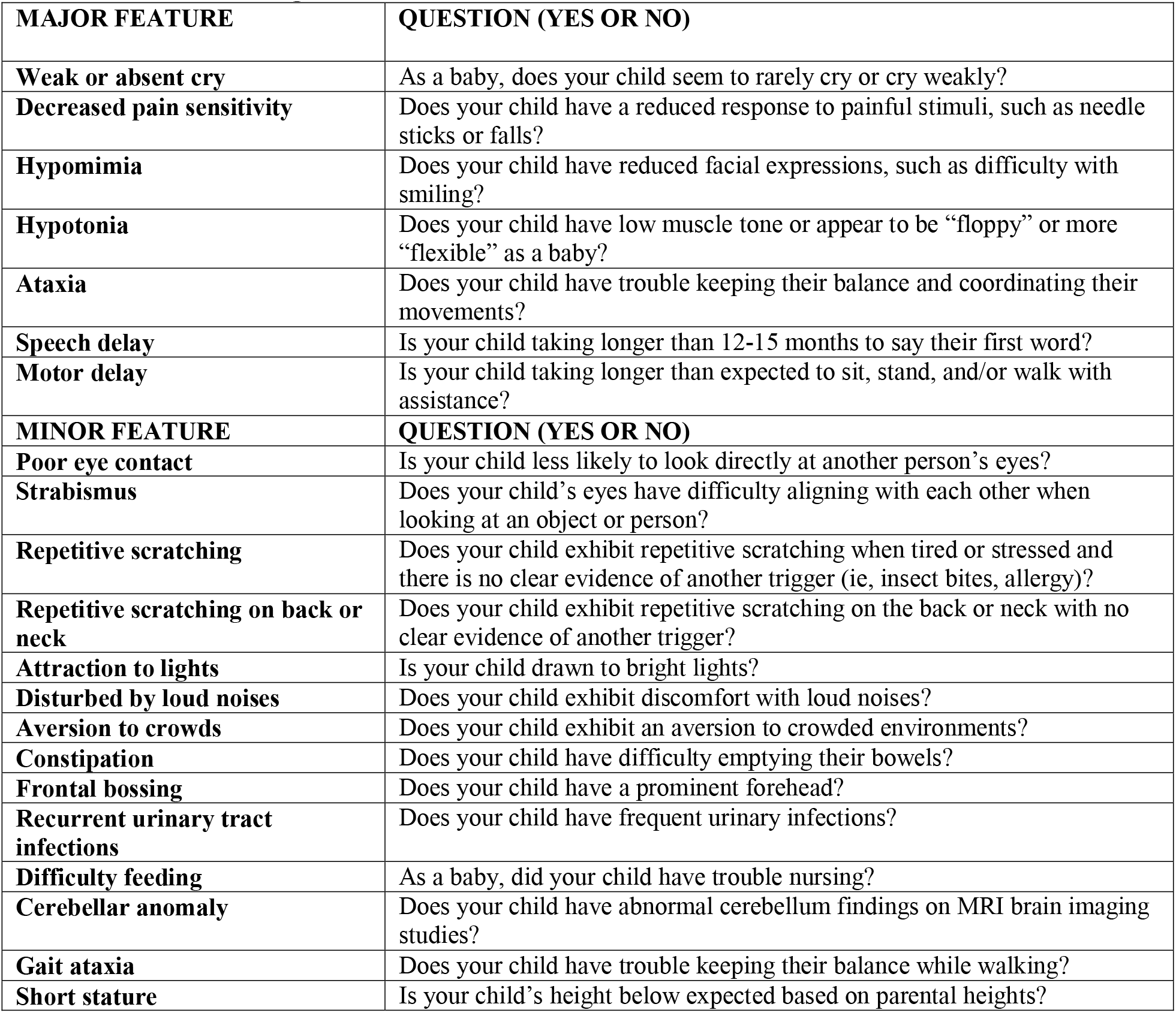
HADDS-Diagnostic Behavioral Scale Questionnaire. The HADDS-DBS scale is comprised of seven major and 14 minor features and designed for use by parents and clinicians. Seven of the clinical features are considered “major” due to their high prevalence among the patients in the current cohort. The remaining 14 clinical features are considered “minor”.

| MAJOR FEATURE | QUESTION (YES OR NO) |
| --- | --- |
| **Weak or absent cry** | As a baby, does your child seem to rarely cry or cry weakly? |
| **Decreased pain sensitivity** | Does your child have a reduced response to painful stimuli, such as needle sticks or falls? |
| **Hypomimia** | Does your child have reduced facial expressions, such as difficulty with smiling? |
| **Hypotonia** | Does your child have low muscle tone or appear to be “floppy” or more “flexible” as a baby? |
| **Ataxia** | Does your child have trouble keeping their balance and coordinating their movements? |
| **Speech delay** | Is your child taking longer than 12-15 months to say their first word? |
| **Motor delay** | Is your child taking longer than expected to sit, stand, and/or walk with assistance? |
| MINOR FEATURE | QUESTION (YES OR NO) |
| **Poor eye contact** | Is your child less likely to look directly at another person’s eyes? |
| **Strabismus** | Does your child’s eyes have difficulty aligning with each other when looking at an object or person? |
| **Repetitive scratching** | Does your child exhibit repetitive scratching when tired or stressed and there is no clear evidence of another trigger (ie, insect bites, allergy)? |
| **Repetitive scratching on back or neck** | Does your child exhibit repetitive scratching on the back or neck with no clear evidence of another trigger? |
| **Attraction to lights** | Is your child drawn to bright lights? |
| **Disturbed by loud noises** | Does your child exhibit discomfort with loud noises? |
| **Aversion to crowds** | Does your child exhibit an aversion to crowded environments? |
| **Constipation** | Does your child have difficulty emptying their bowels? |
| **Frontal bossing** | Does your child have a prominent forehead? |
| **Recurrent urinary tract infections** | Does your child have frequent urinary infections? |
| **Difficulty feeding** | As a baby, did your child have trouble nursing? |
| **Cerebellar anomaly** | Does your child have abnormal cerebellum findings on MRI brain imaging studies? |
| **Gait ataxia** | Does your child have trouble keeping their balance while walking? |
| **Short stature** | Is your child’s height below expected based on parental heights? |

To determine the specificity of the HADDS-DBS, we applied the diagnostic scale to three neurodevelopmental disorders with similar clinical features (hypotonia, delayed development, ataxia, and variable cognitive impairment), but do not have *EBF3* gene alterations (**Table S10**). Three studies were selected that described eight individuals with Harel-Yoon syndrome (HYS [MIM: 617183]) caused by *ATAD3A* variants^38^, 20 individuals with Mental Retardation and Microcephaly with Pontine and Cerebellar Hypoplasia (MICPCH [MIM: 300749]) caused by *CASK* variants^39^, and 15 individuals with Takenouchi-Kosaki syndrome (TKS [MIM: 616737]) caused by *CDC42* variants^40^. Of the eight individuals with HYS^38^, two were classified Tier-1 (25%) and six as Tier-2 (75%). For the 20 individuals with MICPCH^39^, 10 were classified as Tier-1 (50%) and 10 as Tier-2 (50%). Finally, of the 15 individuals with TKS^40^, all were classified as Tier-1 (100%). Statistical comparisons with one-way ANOVA reveal a significant difference for the phenotypic stratification of the HADDS meta-analysis compared to individuals with either HYS, MICPCH, or TKS (**Fig.4A**, *p*<0.051, F=8.34) with sensitivity=0.29, specificity=1.0, PPV=1.0, and NPV=0.64. In contrast, comparison of our HADDS cohort to individuals with either HYS, MICPCH, or TKS reveal sensitivity=0.97, specificity=1.0, PPV=1.0, and NPV=0.97, consistent with the comprehensive phenotypic ascertainment in our study cohort.

### Correlating *EBF3* variant type with severity of motor and language delays

We used the HADDS-DDSS to identify molecular phenotype groups with differential risk of motor and language impairments (**Table S7**). Of the combined 42 individuals diagnosed with HADDS who were more than three years old, we classified seven individuals as Tier-1, 18 as Tier-2, eight as Tier-3, and nine as Tier-4. We examined the HADDS-DDSS classification relative to the *EBF3* variant type (noncoding, gene deletion, or coding) and the location of coding variants relative to the ZNF. For the seven individuals with *EBF3* noncoding variants or gene deletions, one was classified as Tier-1, three as Tier-2, one as Tier-3, and two as Tier-4 severity (**Fig.4B**). For the 21 individuals with *EBF3* coding variants located outside of the ZNF, six were classified as Tier-1, 11 as Tier-2, three as Tier-3, and one as Tier-4. Finally, for the 14 individuals with *EBF3* coding variants located in the ZNF region, the HADDS-DDSS classified four individuals as Tier-2, four as Tier-3, and six as Tier-4 (**Fig.4B**). A significant correlation was identified between increased clinical severity and coding variants within five amino acids of the ZNF (**Fig.4B**, *p*<0.01, F=7.72). Intriguingly, more than 25% of the individuals with HADDS have *EBF3* variants located within the ZNF, which may reflect a selection bias for consistent phenotypes or increased clinical testing due to more pronounced phenotypic features.

## DISCUSSION

Despite increased information available regarding *EBF3*-related HADDS, the need remains for a clear understanding of the phenotypic variability and how the *EBF3* genotype affects specific clinical findings. Our comprehensive phenotyping in a large cohort of individuals molecularly diagnosed with pathogenic *EBF3* variants allow for an expanded understanding of the variation in clinical outcomes and improved clinical management (**Fig.S1**). Neuroanatomical studies reveal that the cerebellum is the most common structure perturbed in individuals with *EBF3* pathogenic variants. Extensive anatomical and functional studies revealed critical roles for the cerebellum in motor and non-motor functions, including language, cognition, and social behavior^37^. Thus, many of the features observed in individuals with HADDS may be consistent with cerebellar dysfunction. To further understand the relationship between the *EBF3* molecular phenotype and clinical outcome, we designed two assessment scales that determine an individual’s risk for having a pathogenic *EBF3* variant (HADDS-DBS) and their degree of functional impairments (HADDS-DDSS). Our findings validated the clinical utility of the scales, revealed critical insights into the *EBF3* genotype-phenotype map, and demonstrated that molecular phenotypes correlate with quantitative clinical findings (**Fig.4**).

The ability to broadly predict the risk of a pathogenic *EBF3* variant advances clinical diagnosis, facilitates prognosis, and aids in resolving *EBF3* variants of uncertain significance (VUS). Moreover, we were interested in determining whether the *EBF3* molecular phenotype could provide insights into the variation of clinical outcomes. By integrating the HADDS-DDSS assessment with the *EBF3* molecular phenotype, we found that missense variants within the ZNF define a subgroup of individuals at risk for persistent motor and language impairments. This finding may be related to a stronger dominant-negative effect resulting from *EBF3* missense variants located affecting the ZNF. Haploinsufficient non-coding *EBF3* variants resulting in nonsense mediated decay or *EBF3* gene deletion appears to correlate with intermediate clinical outcomes. However, this interpretation is limited by the small sample size. Furthermore, the variation in clinical outcomes associated with *EBF3* variants raises the possibility that subtle impairments in EBF3 function or protein levels may result in a limited subset of HADDS features or manifest primarily as a non-syndromic ASD or non-specific developmental delay. This hypothesis is supported by prior work identifying *EBF3* as one of ∼1,000 genes with significant enrichment of *de novo* loss-of-function variants in individuals with ASD and DD/ID^26^.

By using the largest set of clinically annotated *EBF3* variants examined to date, we strengthen prior findings by showing that HADDS-associated *EBF3* variants are related to a wide spectrum of clinical outcomes, identified Mendelian transmission of potentially hypomorphic variants, and expanded our understanding of cardinal diagnostic features. Furthermore, we found that the distribution of pathogenic *EBF3* variants remains predominantly in the DNA-binding domain, suggesting that variants in the C-terminal domains are less damaging to EBF3 function. Of note, we found that *EBF3* variants affecting the ZNF are associated with an increased risk of persistent motor and/or language impairments. These differential risk profiles may account for the increased frequency and severity of *de novo* missense variants in the ZNF and surrounding nucleotides, whereas milder variants resulting in subtle impairments in EBF3 function may not be identified primarily on the basis of a clinical HADDS diagnosis.

Despite advances in our recognition of *EBF3*-related HADDS, the neurobiological basis of differential risk profiles based on *EBF3* variant type and location remains to be elucidated. As EBF3 functions through homo- or heterodimerization, the missense variants may result in a dominant-negative effect that alters the function of the wildtype EBF3 protein and reduces the overall amount of functional EBF3 dimers. This would result in increased perturbations to the EBF3-dependent transcriptional regulation in the developing nervous system. In contrast, *EBF3* single gene deletion or non-coding variants may preserve functional EBF3 dimers but result in an overall reduction of EBF3 available to regulate developmental processes. Ideally, a comprehensive analysis would include the effect of variation on EBF3’s protein-protein interactions, subcellular localizations, regulation of target gene expression profiles, and nuclear function. The increasing recognition of individuals at risk for pathogenic *EBF3* variants and new high-throughput assays might make such datasets available for expanded analysis of the neurobiological consequences of the molecular phenotype.

In conclusion, we show that pathogenic *EBF3* variants cause a broad spectrum of phenotypic severity. This expands our understanding of cardinal clinical features in HADDS. Through the integration of the HADDS-DBS and HADDS-DDSS assessments with the molecular phenotype, we found that a subset of individuals with *EBF3* missense variants affecting the ZNF have a higher risk of persistent functional impairments. Furthermore, the phenotypic overlap between HADDS with ASD and DD/ID suggest that subtle EBF3 dysfunction may result in non-syndromic features and that HADDS may share common pathogenic mechanisms with ASD and DD/ID. Longitudinal follow-up of these individuals in conjunction with new prospective recruitment efforts will be needed to comprehensively delineate the *EBF3* genotype-phenotype map, which may lead to promising avenues for therapeutic interventions.

## Data Availability

De-identified data regarding molecular and clinical findings are available upon request.

## DECLARATION OF INTERESTS

The authors declare no competing interests.

### ACKNOWLEDGEMENTS

We thank the families for participation in this study, Keri Ramsey with clinical assistance, and Mingshan Xue, Sahana Murthy, Harim Delgado-Seo, Maimuna Paul, and Joshi Stephen for manuscript feedback. There is no targeted funding for this study. H.T.C. is partially supported from NIH-1DP5OD026428. C.D. is supported by the BCM Medical Scientist Training Program. N.D.B. was supported by the Autism Science Foundation. V.L. is supported by The Gordon and Mary Cain Foundation and Annie and Bob Graham. EB and GZ are members of the European Reference Network for Rare Neurological Diseases-Project #739510. EB, EMV, and GZ are supported by the Ministry of Health (Ricerca Corrente 2020, Ricerca Finalizzata NET-2013-02356160).

## WEB RESOURCES

OMIM, https://www.omim.org

GenBank, https://www.ncbi.nlm.nih.gov/genbank

Centers for Disease Control and Prevention growth charts, https://www.cdc.gov/growthcharts/index.htm

World Health Organization growth charts, https://www.who.int/childgrowth/standards/en/ gnomAD-Genome Aggregation Database, https://gnomad.broadinstitute.org/

CADD-Combined Annotation Dependent Depletion, https://cadd.gs.washington.edu/snv UCSC Genome Browser (GRCh37/hg19), https://genome.ucsc.edu/cgi-bin/hgGateway

## DATA AND CODE AVAILABILITY

The de-identified data supporting the current study have not been deposited in a public repository but are available from the corresponding author on request.

## REFERENCES

1. Decker, T., Pasca di Magliano, M., McManus, S., Sun, Q., Bonifer, C., Tagoh, H., and Busslinger, M. (2009). Stepwise activation of enhancer and promoter regions of the B cell commitment gene Pax5 in early lymphopoiesis. Immunity 30, 508–520.

2. Maier, H., Ostraat, R., Gao, H., Fields, S., Shinton, S.A., Medina, K.L., Ikawa, T., Murre, C., Singh, H., Hardy, R.R., et al. (2004). Early B cell factor cooperates with Runx1 and mediates epigenetic changes associated with mb-1 transcription. Nat. Immunol. 5, 1069–1077.

3. Treiber, T., Mandel, E.M., Pott, S., Györy, I., Firner, S., Liu, E.T., and Grosschedl, R. (2010). Early B cell factor 1 regulates B cell gene networks by activation, repression, and transcription-independent poising of chromatin. Immunity 32, 714–725.

4. Green, Y.S., and Vetter, M.L. (2011). EBF proteins participate in transcriptional regulation of Xenopus muscle development. Dev. Biol. 358, 240–250.

5. Kuriki, M., Sato, F., Arai, H.N., Sogabe, M., Kaneko, M., Kiyonari, H., Kawakami, K., Yoshimoto, Y., Shukunami, C., and Sehara-Fujisawa, A. (2020). Transient and lineage-restricted requirement of Ebf3 for sternum ossification. Development 147,.

6. Wang, S.S., Lewcock, J.W., Feinstein, P., Mombaerts, P., and Reed, R.R. (2004). Genetic disruptions of O/E2 and O/E3 genes reveal involvement in olfactory receptor neuron projection. Development 131, 1377–1388.

7. Sugiyama, T., Omatsu, Y., and Nagasawa, T. (2019). Niches for hematopoietic stem cells and immune cell progenitors. Int. Immunol. 31, 5–11.

8. Liberg, D., Sigvardsson, M., and Akerblad, P. (2002). The EBF/Olf/Collier family of transcription factors: regulators of differentiation in cells originating from all three embryonal germ layers. Mol. Cell. Biol. 22, 8389–8397.

9. Chiara, F., Badaloni, A., Croci, L., Yeh, M.L., Cariboni, A., Hoerder-Suabedissen, A., Consalez, G.G., Eickholt, B., Shimogori, T., Parnavelas, J.G., et al. (2012). Early B-cell factors 2 and 3 (EBF2/3) regulate early migration of Cajal-Retzius cells from the cortical hem. Dev. Biol. 365, 277–289.

10. Fulp, C.T., Cho, G., Marsh, E.D., Nasrallah, I.M., Labosky, P.A., and Golden, J.A. (2008). Identification of Arx transcriptional targets in the developing basal forebrain. Hum. Mol. Genet. 17, 3740–3760.

11. Garel, S., Garcia-Dominguez, M., and Charnay, P. (2000). Control of the migratory pathway of facial branchiomotor neurones. Development 127, 5297–5307.

12. Harms, F.L., Girisha, K.M., Hardigan, A.A., Kortüm, F., Shukla, A., Alawi, M., Dalal, A., Brady, L., Tarnopolsky, M., Bird, L.M., et al. (2017). Mutations in EBF3 Disturb Transcriptional Profiles and Cause Intellectual Disability, Ataxia, and Facial Dysmorphism. Am. J. Hum. Genet. 100, 117–127.

13. Lewandowski, R.C., Kukolich, M.K., Sears, J.W., and Mankinen, C.B. (1978). Partial deletion 10q. Hum Genet 42, 339–343.

14. Shapiro, S.D., Hansen, K.L., Pasztor, L.M., DiLiberti, J.H., Jorgenson, R.J., Young, R.S., and Moore, C.M. (1985). Deletions of the long arm of chromosome 10. Am J Med Genet 20, 181– 196.

15. Mehta, L., Duckett, D.P., and Young, I.D. (1987). Behaviour disorder in monosomy 10qter. J. Med. Genet. 24, 185–186.

16. Tanabe, S., Akiba, T., Katoh, M., and Satoh, T. (1999). Terminal deletion of chromosome 10q: clinical features and literature review. Pediatr Int 41, 565–567.

17. Irving, M., Hanson, H., Turnpenny, P., Brewer, C., Ogilvie, C.M., Davies, A., and Berg, J. (2003). Deletion of the distal long arm of chromosome 10; is there a characteristic phenotype? A report of 15 de novo and familial cases. Am. J. Med. Genet. A 123A, 153–163.

18. Chao, H.-T., Davids, M., Burke, E., Pappas, J.G., Rosenfeld, J.A., McCarty, A.J., Davis, T., Wolfe, L., Toro, C., Tifft, C., et al. (2017). A Syndromic Neurodevelopmental Disorder Caused by De Novo Variants in EBF3. Am J Hum Genet 100, 128–137.

19. Sleven, H., Welsh, S.J., Yu, J., Churchill, M.E.A., Wright, C.F., Henderson, A., Horvath, R., Rankin, J., Vogt, J., Magee, A., et al. (2017). De Novo Mutations in EBF3 Cause a Neurodevelopmental Syndrome. Am. J. Hum. Genet. 100, 138–150.

20. Lopes, F., Soares, G., Gonçalves-Rocha, M., Pinto-Basto, J., and Maciel, P. (2017). Whole Gene Deletion of EBF3 Supporting Haploinsufficiency of This Gene as a Mechanism of Neurodevelopmental Disease. Front Genet 8, 143.

21. Blackburn, P.R., Barnett, S.S., Zimmermann, M.T., Cousin, M.A., Kaiwar, C., Pinto E Vairo, F., Niu, Z., Ferber, M.J., Urrutia, R.A., Selcen, D., et al. (2017). Novel de novo variant in EBF3 is likely to impact DNA binding in a patient with a neurodevelopmental disorder and expanded phenotypes: patient report, in silico functional assessment, and review of published cases. Cold Spring Harb Mol Case Stud 3, a001743.

22. Tanaka, A.J., Cho, M.T., Willaert, R., Retterer, K., Zarate, Y.A., Bosanko, K., Stefans, V., Oishi, K., Williamson, A., Wilson, G.N., et al. (2017). De novo variants in EBF3 are associated with hypotonia, developmental delay, intellectual disability, and autism. Cold Spring Harb Mol Case Stud 3,.

23. Beecroft, S.J., Olive, M., Quereda, L.G., Gallano, M.P., Ojanguren, I., McLean, C., McCombe, P., Laing, N.G., and Ravenscroft, G. (2019). Cylindrical spirals in two families: Clinical and genetic investigations. Neuromuscul. Disord.

24. Harkness, J.R., Beaman, G.M., Teik, K.W., Sidhu, S., Sayer, J.A., Cordell, H.J., Thomas, H.B., Wood, K., Stuart, H.M., Woolf, A.S., et al. (2020). Early B-cell Factor 3-Related Genetic Disease Can Mimic Urofacial Syndrome. Kidney Int Rep 5, 1823–1827.

25. Hildebrand, M.S., Jackson, V.E., Scerri, T.S., Van Reyk, O., Coleman, M., Braden, R.O., Turner, S., Rigbye, K.A., Boys, A., Barton, S., et al. (2020). Severe childhood speech disorder. Neurology 94, e2148.

26. Samocha, K.E., Robinson, E.B., Sanders, S.J., Stevens, C., Sabo, A., McGrath, L.M., Kosmicki, J.A., Rehnström, K., Mallick, S., Kirby, A., et al. (2014). A framework for the interpretation of de novo mutation in human disease. Nat. Genet. 46, 944–950.

27. Flegal, K.M., and Cole, T.J. (2013). Construction of LMS parameters for the Centers for Disease Control and Prevention 2000 growth charts (US Department of Health and Human Services, Centers for Disease Control and …).

28. Kuczmarski, R.J. (2002). 2000 CDC Growth Charts for the United States: methods and development (Department of Health and Human Services, Centers for Disease Control and …).

29. World Health Organization (2007). WHO child growth standards: methods and development (World Health Organization).

30. Milestone Checklist.

31. Hagman, J., Gutch, M.J., Lin, H., and Grosschedl, R. (1995). EBF contains a novel zinc coordination motif and multiple dimerization and transcriptional activation domains. EMBO J. 14, 2907–2916.

32. Magewu, A.N., and Jones, P.A. (1994). Ubiquitous and tenacious methylation of the CpG site in codon 248 of the p53 gene may explain its frequent appearance as a mutational hot spot in human cancer. Mol. Cell. Biol. 14, 4225–4232.

33. Mancini, D., Singh, S., Ainsworth, P., and Rodenhiser, D. (1997). Constitutively methylated CpG dinucleotides as mutation hot spots in the retinoblastoma gene (RB1). Am. J. Hum. Genet. 61, 80–87.

34. Goriely, A., McGrath, J.J., Hultman, C.M., Wilkie, A.O.M., and Malaspina, D. (2013). “Selfish spermatogonial selection”: a novel mechanism for the association between advanced paternal age and neurodevelopmental disorders. Am J Psychiatry 170, 599–608.

35. Dhokarh, D., and Abyzov, A. (2016). Elevated variant density around SV breakpoints in germline lineage lends support to error-prone replication hypothesis. Genome Res 26, 874–881.

36. Bolceková, E., Mojzeš, M., Van Tran, Q., Kukal, J., Ostrý, S., Kulišťák, P., and Rusina, R. (2017). Cognitive impairment in cerebellar lesions: a logit model based on neuropsychological testing. Cerebellum Ataxias 4, 13.

37. Strick, P.L., Dum, R.P., and Fiez, J.A. (2009). Cerebellum and nonmotor function. Annu. Rev. Neurosci. 32, 413–434.

38. Harel, T., Yoon, W.H., Garone, C., Gu, S., Coban-Akdemir, Z., Eldomery, M.K., Posey, J.E., Jhangiani, S.N., Rosenfeld, J.A., Cho, M.T., et al. (2016). Recurrent De Novo and Biallelic Variation of ATAD3A, Encoding a Mitochondrial Membrane Protein, Results in Distinct Neurological Syndromes. Am. J. Hum. Genet. 99, 831–845.

39. Moog, U., Kutsche, K., Kortüm, F., Chilian, B., Bierhals, T., Apeshiotis, N., Balg, S., Chassaing, N., Coubes, C., Das, S., et al. (2011). Phenotypic spectrum associated with CASK loss-of-function mutations. J. Med. Genet. 48, 741–751.

40. Martinelli, S., Krumbach, O.H.F., Pantaleoni, F., Coppola, S., Amin, E., Pannone, L., Nouri, K., Farina, L., Dvorsky, R., Lepri, F., et al. (2018). Functional Dysregulation of CDC42 Causes Diverse Developmental Phenotypes. Am. J. Hum. Genet. 102, 309–320.

