## Supplemental Materials for "Integrated phenotypic and mutational approach defines EBF3-related HADD syndrome genotype-phenotype relationships" (from another version of this preprint: version 1, 2020-12-08)

**Web Resources**

OMIM, https://www.omim.org

GenBank, https://www.ncbi.nlm.nih.gov/genbank

Centers for Disease Control and Prevention growth charts, https://www.cdc.gov/growthcharts/index.htm

World Health Organization growth charts, https://www.who.int/childgrowth/standards/en/

gnomAD-Genome Aggregation Database, https://gnomad.broadinstitute.org/

CADD-Combined Annotation Dependent Depletion, https://cadd.gs.washington.edu/snv

UCSC Genome Browser (GRCh37/hg19), https://genome.ucsc.edu/cgi-bin/hgGateway

**Table S1: Summary of clinical findings in cohort of individuals with *EBF3*-related HADDS**

General features, molecular phenotype, and clinical findings identified through comprehensive phenotypic ascertainment in 33 individuals molecularly diagnosed with *EBF3*-related HADDS and enrolled in the study. Abbreviations: year (yr), month (mo), not available (n/a), World Health Organization (WHO), occipito-frontal circumference (OFC), National Center for Biotechnology Information (NCBI), copy number variant (CNV), electroencephalogram (EEG), attention deficit hyperactivity disorder (ADHD), magnetic resonance imaging (MRI), urinary tract infection (UTI)

**Table S2: Summary of clinical findings in meta-analysis of published individuals with *EBF3*-related HADDS**

General features, molecular phenotype, and clinical findings identified through comprehensive phenotypic ascertainment in 34 individuals molecularly diagnosed with *EBF3*-related HADDS and previously reported in the literature (PMIDs: 28017372, 28017373, 28017370, 29162653, 29062322, 28487885, 31952901, 32345733, 33102976). Abbreviations: year (yr), month (mo), not available (n/a), reported within normal limits (wnl), World Health Organization (WHO), occipito-frontal circumference (OFC), National Center for Biotechnology Information (NCBI), copy number variant (CNV), electroencephalogram (EEG), attention deficit hyperactivity disorder (ADHD), magnetic resonance imaging (MRI), urinary tract infection (UTI)

**Table S3: Growth parameters of individuals with *EBF3*-related HADDS**

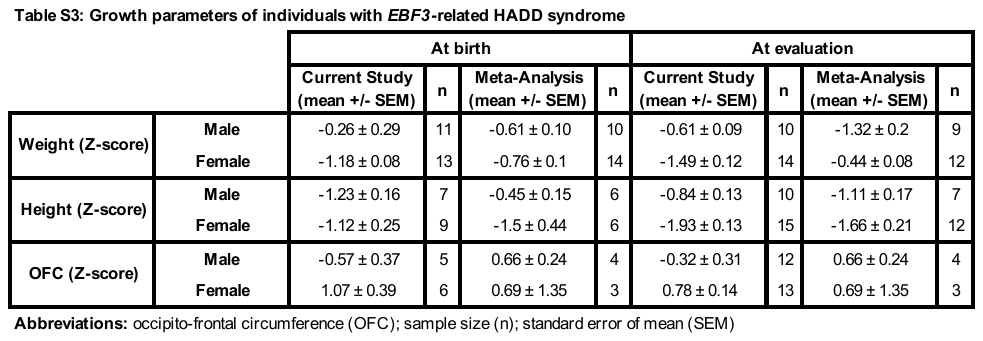

Comparison of height, weight, and occipito-frontal circumference (OFC) growth parameters in individuals enrolled in our study and in the meta-analysis. LMS statistics provided by the CDC and WHO were used to estimate the z-scores for growth parameters at birth and most recent evaluation for males and females. Data shown as mean ± SEM. Sample size (n) shown in table.

**Table S4: Summary of developmental milestones in cohort of individuals with *EBF3*-related HADDS**

Gross motor and language milestones identified through comprehensive phenotypic ascertainment in 33 individuals molecularly diagnosed with *EBF3*-related HADDS and enrolled in the study. Abbreviations: year (yr), month (mo), not available (n/a)

**Table S5: Summary of developmental milestones in meta-analysis of published individuals with *EBF3*-related HADDS**

Gross motor and language milestones identified through comprehensive phenotypic ascertainment in 34 individuals molecularly diagnosed with *EBF3*-related HADDS and previously reported in the literature (PMIDs: 28017372, 28017373, 28017370, 29162653, 29062322, 28487885, 31952901, 32345733, 33102976). Abbreviations: year (yr), month (mo), not available (n/a)

**Table S6: HADDS-Diagnostic Behavioral Scale (HADDS-DBS) tier assignments**

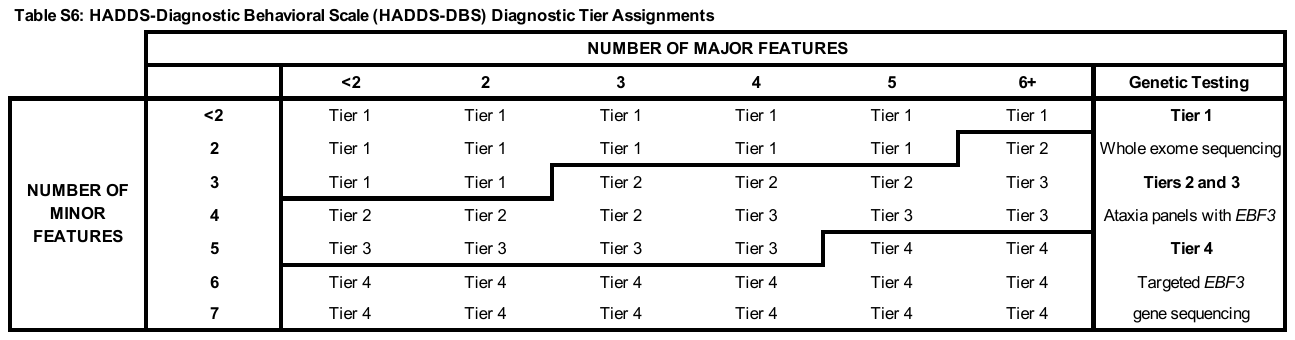

The HADDS-DBS is comprised of a four-tier distribution scale (Tiers-1 through -4). Based on the number of major and minor diagnostic features identified, the individuals are phenotypically stratified into four groups, with Tier-1 indicating a low risk and Tier-4 indicating a high risk for a pathogenic *EBF3* variant and co-morbidities. For Tiers-1 and -2, the HADDS-DBS recommends broader genetic testing, as the phenotypic findings are either not suggestive of HADDS or there is insufficient information for phenotypic stratification. For Tier-3, there is a moderate risk for a pathogenic *EBF3* variant, and ataxia syndrome gene panels that include *EBF3* are recommended for molecular diagnostics. A targeted *EBF3* gene test is recommended for Tier-4 individuals due to a higher risk of pathogenic *EBF3* variants.

**Table S7: HADDS-Developmental Delay Severity Scale (HADDS-DDSS) tier assignments**

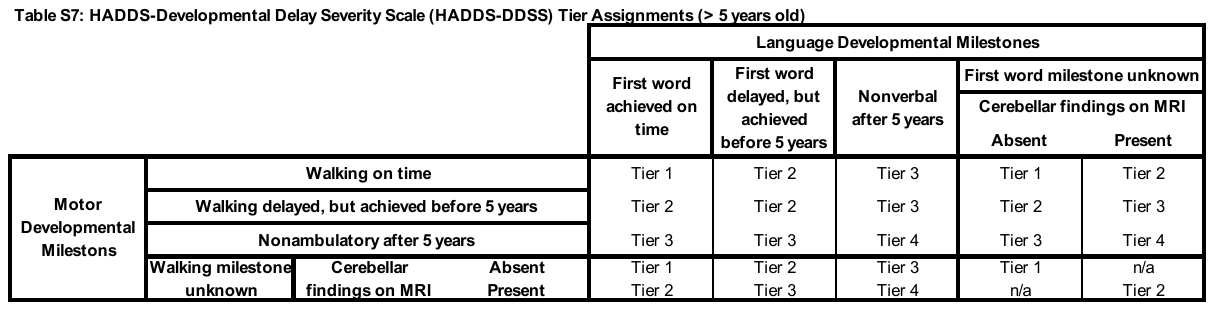

The HADDS Developmental Delay Severity Scale (HADDS-DDSS) is comprised of four tiers based on the magnitude of delayed development and neuroanatomical findings for prognostication and genotype-phenotype mapping. A higher-numbered tier indicates increasingly delayed development, persistent limitations in motor and language abilities, and presence of cerebellar alterations on MRI studies.

**Table S8: Summary of HADDS-DBS and HADDS-DDSS tier classifications in cohort of individuals with *EBF3*-related HADDS**

Summary of tier assignments per proband in the study cohort.

**Table S9: Summary of HADDS-DBS and HADDS-DDSS tier classifications in meta-analysis of individuals with *EBF3*-related HADDS**

Summary of tier assignments per proband in meta-analysis.

**Figure S1: HADDS phenotypic infographic tool for patients, families, and clinicians**

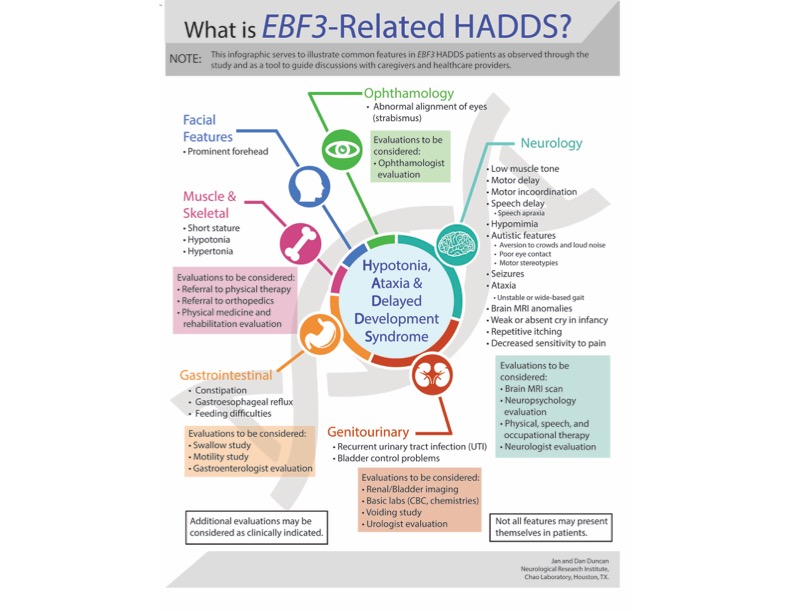

An infographic summarizing the most commonly reported symptoms of HADDS. Findings occurring in more than 30% of individuals are shown with suggested clinical evaluations.
